# Viable SARS-CoV-2 in the air of a hospital room with COVID-19 patients

**DOI:** 10.1101/2020.08.03.20167395

**Authors:** John A. Lednicky, Michael Lauzardo, Z. Hugh Fan, Antarpreet Jutla, Trevor B. Tilly, Mayank Gangwar, Moiz Usmani, Sripriya Nannu Shankar, Karim Mohamed, Arantza Eiguren-Fernandez, Caroline J. Stephenson, Md. Mahbubul Alam, Maha A. Elbadry, Julia C. Loeb, Kuttinchantran Subramaniam, Thomas B. Waltzek, Kartikeya Cherabuddi, J. Glenn Morris, Chang-Yu Wu

**Affiliations:** Department of Environmental and Global Health, College of Public Health and Health Professions, University of Florida, USA; Emerging Pathogens Institute, University of Florida, USA; Division of Infectious Diseases and Global Medicine, Department of Medicine, College of Medicine, University of Florida, USA; Department of Mechanical & Aerospace Engineering, College of Engineering, University of Florida, USA; J. Crayton Pruitt Family Department of Biomedical Engineering, University of Florida, USA; Department of Environmental Engineering Sciences, College of Engineering, University of Florida, USA; Aerosol Dynamics Inc., Berkeley, California, USA; Department of Infectious Diseases and Immunology, College of Veterinary Medicine, University of Florida, USA

## Abstract

**Background:** There currently is substantial controversy about the role played by SARS-CoV-2 in aerosols in disease transmission, due in part to detections of viral RNA but failures to isolate viable virus from clinically generated aerosols.

**Methods:** Air samples were collected in the room of two COVID-19 patients, one of whom had an active respiratory infection with a nasopharyngeal (NP) swab positive for SARS-CoV-2 by RT-qPCR. By using VIVAS air samplers that operate on a gentle water-vapor condensation principle, material was collected from room air and subjected to RT-qPCR and virus culture. The genomes of the SARS-CoV-2 collected from the air and of virus isolated in cell culture from air sampling and from a NP swab from a newly admitted patient in the room were sequenced.

**Findings:** Viable virus was isolated from air samples collected 2 to 4.8m away from the patients. The genome sequence of the SARS-CoV-2 strain isolated from the material collected by the air samplers was identical to that isolated from the NP swab from the patient with an active infection. Estimates of viable viral concentrations ranged from 6 to 74 TCID_50_ units/L of air.

**Interpretation:** Patients with respiratory manifestations of COVID-19 produce aerosols in the absence of aerosol-generating procedures that contain viable SARS-CoV-2, and these aerosols may serve as a source of transmission of the virus.

**Funding:** Partly funded by Grant No. 2030844 from the National Science Foundation and by award 1R43ES030649 from the National Institute of Environmental Health Sciences of the National Institutes of Health, and by funds made available by the University of Florida Emerging Pathogens Institute and the Office of the Dean, University of Florida College of Medicine.

**Research in context:** *Evidence before this study:* Various studies report detection of SARS-CoV-2 in material collected by air samplers positioned in clinics and in some public spaces. For those studies, detection of SARS-CoV-2 has been by indirect means; instead of virus isolation, the presence of the virus in material collected by air samplers has been through RT-PCR detection of SARS-CoV-2 RNA. However, questions have been raised about the clinical significance of detection of SARS-CoV-2 RNA, particularly as airborne viruses are often inactivated by exposure to UV light, drying, and other environmental conditions, and inactivated SARS-CoV-2 cannot cause COVID-19.

*Added value of this study:* Our virus isolation work provides direct evidence that SARS-CoV-2 in aerosols can be viable and thus pose a risk for transmission of the virus. Furthermore, we show a clear progression of virus-induced cytopathic effects in cell culture, and demonstrate that the recovered virus can be serially propagated. Moreover, we demonstrate an essential link: the viruses we isolated in material collected in four air sampling runs and the virus in a newly admitted symptomatic patient in the room were identical. These findings strengthen the notion that airborne transmission of viable SARS-CoV-2 is likely and plays a critical role in the spread of COVID-19.

*Implications of all the available evidence:* Scientific information on the mode of transmission should guide best practices Current best practices for limiting the spread of COVID-19. Transmission secondary to aerosols, without the need for an aerosol-generating procedure, especially in closed spaces and gatherings, has been epidemiologically linked to exposures and outbreaks. For aerosol-based transmission, measures such as physical distancing by 6 feet would not be helpful in an indoor setting and would provide a false-sense of security. With the current surges of cases, to help stem the COVID-19 pandemic, clear guidance on control measures against SARS-CoV-2 aerosols are needed.

## Introduction

Severe acute respiratory syndrome coronavirus 2 (SARS-CoV-2), genus *Betacoronavirus*, subgenus *Sarbecovirus*, family *Coronaviridae*, is a positive-polarity single-stranded RNA virus that probably originated in bats^1–3^ and is the causative agent of coronavirus disease of 2019 (COVID-19).^4^ The dynamics of the COVID-19 pandemic have proven to be complex. Many challenges remain pertaining to a better understanding of the epidemiology, pathology, and transmission of COVID-19. For example, the clinical manifestations of COVID-19 range from an asymptomatic infection, mild respiratory illness to pneumonia, respiratory failure, multi-organ failure, and death.^5-7^ Diarrhea due to gastro-intestinal infection can also occur, and *in vitro* modeling suggests that the virus infects human gut enterocytes.^8^ SARS-CoV-2 RNA can be found in rectal swabs and fecal aerosols, even after nasal-pharyngeal testing has turned negative,^9-12^ suggesting that a fecal–oral transmission route may be possible.

To-date, there has been a strong emphasis on the role of respiratory droplets and fomites in the transmission of SARS-CoV-2.^13,14^ Yet SARS-CoV-2 does not appear to be exclusively inhaled as a droplet, and epidemiologic data are consistent with aerosol transmission of SARS-CoV-2.^15-19^ Furthermore, SARS-CoV-2 genomic RNA has been detected in airborne material collected by air samplers positioned distal to COVID-19 patients.^9, 20-23^ Any respiratory virus that can survive aerosolization poses an inhalation biohazard risk, and van Doremalen *et al*.^24^ experimentally generated aerosol particles with SARS-CoV-2 and found that the virus remained viable during a three-hour testing period. More recently, Fears *et al*.^25^ reported that the virus retained infectivity and integrity for up to 16 hours in laboratory-created respirable-sized aerosols. Nevertheless, finding virus RNA in material collected by an air sampler may not correlate with risk. Indeed, the air we breathe is full of viruses (animal, plant, bacterial, human, etc.), yet a large proportion of the viruses in air are non-viable due to UV-inactivation, drying, etc., and non-viable viruses cannot cause illnesses. Because efforts to isolate virus in cell cultures in the aforementioned air sampling studies in hospital wards were not made,^20,22^ or failed when they were attempted due to overgrowth by faster replicating respiratory viruses,^23^ or provided weak evidence of virus isolation,^21^ uncertainties about the role of aerosols in COVID-19 transmission remain.

It is well known that virus particles collected by various air samplers become inactivated during the air sampling process,^26^ and if such is the case for SARS-CoV-2, this partly explains why it has been difficult to prove that SARS-CoV-2 collected from aerosols is viable. Because we previously collected SARS-CoV-2 from the air of a respiratory illness ward within a clinic but were unable to isolate the virus in cell cultures due to out-competition by other respiratory viruses,^23^ we sought to perform air sampling tests in a hospital room reserved for COVID-19 patients, to lessen the probability of collecting other airborne human respiratory viruses. We thus collected aerosols containing SARS-CoV-2 in a room housing COVID-19 patients using our VIVAS air samplers that collect virus particles without damaging them, thus conserving their viability. These samplers operate using a water-vapor condensation mechanism.^27,28^ Air samplings were performed at the University of Florida Health (UF Health) Shands Hospital, which is a 1,050-bed teaching hospital situated in Gainesville, Florida. As of 10 July 2020, > 200 patients have been treated at the hospital for COVID-19. The current study was conducted as part of ongoing environmental investigations by the UF Health infection control group to assess possible healthcare worker exposure to SARS-CoV-2.

## Methods

Detailed methods are provided in a Technical Appendix. An abbreviated summary of methods is provided below:

### Institutional Review Board (IRB) approval and patients

The study protocol was approved by the UF IRB (study IRB202002102). Patient 1 was a person with coronary artery disease and other co-morbidities who had been transferred from a long-term care facility for COVID-19 treatment the evening before our air sampling tests were initiated; he had a positive NP swab test on admission that was positive for SARS-CoV-2 by RT-PCR. Patient 2 had been admitted four days before the air sampling tests with a mid-brain stroke; the patient had a positive NP swab test for SARS-CoV-2 on admission, but a repeat test was negative, and the patient was in the process of being discharged at the time the air sampling was being done.

### Hospital room

Air samples were collected in a room that was part of a designated COVID-19 ward (Figure 1). The room had six air changes per hour and the exhaust air underwent triple filter treatment (minimum efficiency reporting value [MERV] 14, 75%-85% efficiency for 0.3 µm particles), coil condensation (to remove moisture), and UV-C irradiation prior to recycling 90% of the treated air back to the room.

**Figure 1.**
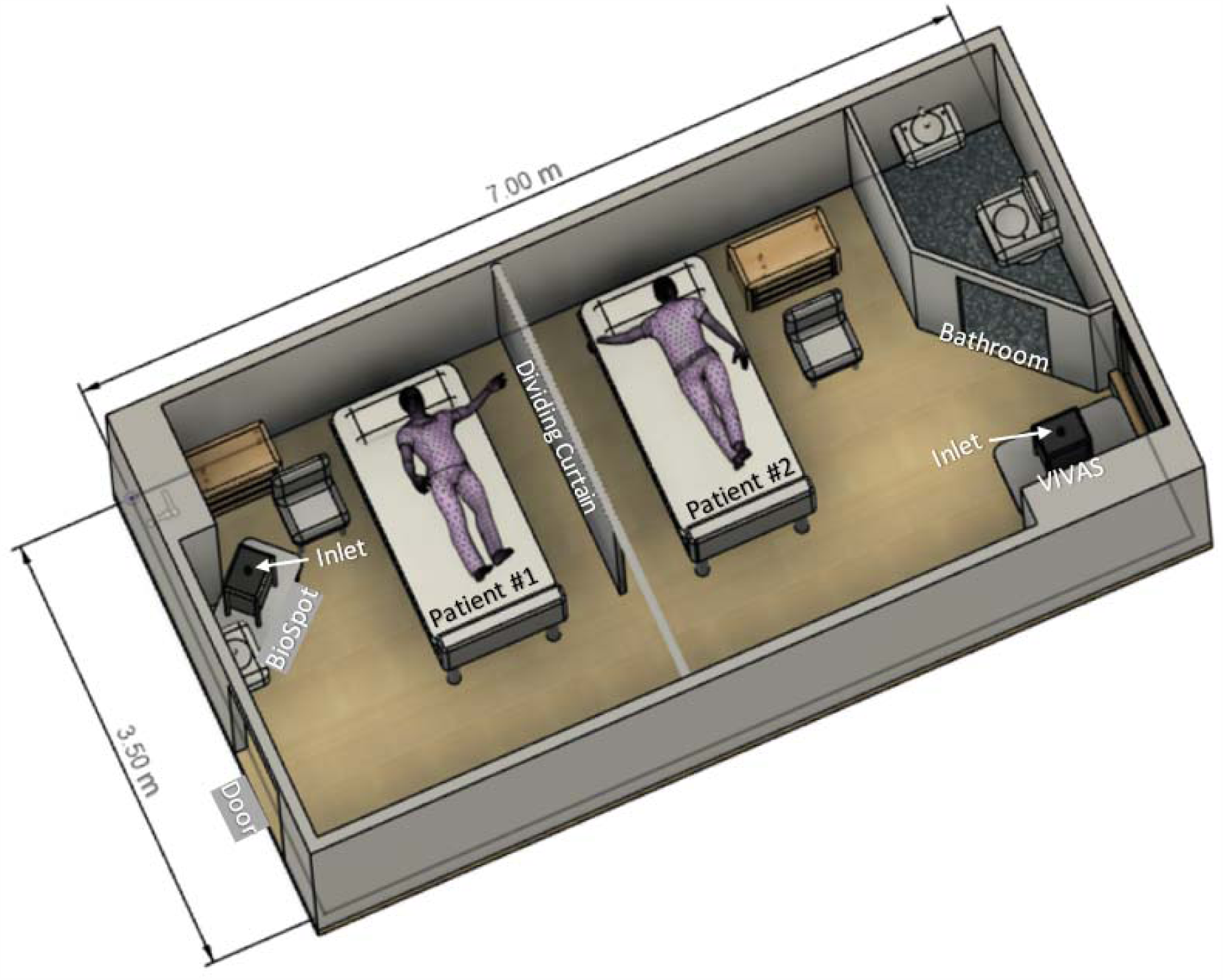
Schematic diagram of room with depiction of patient bed and air-sampler locations.

### Air samplers and sampling parameters

Three serial 3-hr air samplings were performed using our prototype VIVAS air sampler,^23, 27, 28^ as well as a BioSpot-VIVAS BSS300P, which is a commercial version of the VIVAS (available from Aerosol Devices Inc., Ft. Collins, CO). These samplers collect airborne particles using a water-vapor condensation method.^23, 27, 28^ Two samplers were used so that air could be collected/sampled at different sites of the same room during a given air sampling period. For each sampler, the second of the three samplings was performed with a high efficiency particulate arrestance (HEPA) filter affixed to the inlet tube, a process we implement to reveal whether virus detected in consecutive samplings reflect true collection and not detection of residual virus within the collector. The air-samplers were stationed from 2 to 4.8 m away from the patients (Figure 1).

### Detection of SARS-CoV-2 genomic RNA (vRNA) in collection media

vRNA was extracted from virions in collection media and purified by using a QIAamp Viral RNA Mini Kit (Qiagen, Valencia, CA, USA). Twenty-five µL (final volume) real-time reverse-transcription polymerase chain reaction (rtRT-PCR) tests were performed in a BioRad CFX96 Touch Real-Time PCR Detection System using 5 µL of purified vRNA and rtRT-PCR primers and the probe listed in Table 1 that detect a section of the SARS-CoV-2 N-gene.^23^ The N-gene rRT-PCR assay that was used was part of a dual (N- and RdRp-gene) rRT-PCR assay designed by J. Lednicky and does not detect common human alpha- or beta-coronaviruses. Using this particular N-gene rRT-PCR detection system, the limit of detection is about 1.5 SARS-CoV-2 genome equivalents per 25 µL rRT-PCR assay.

**Table 1.**
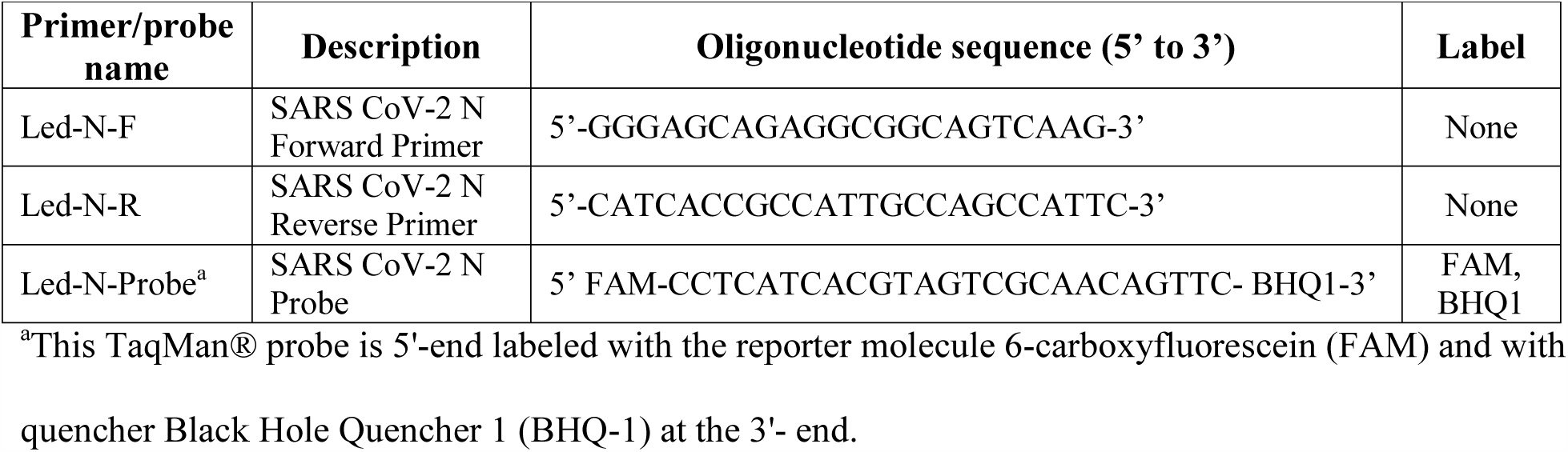
SARS-CoV-2 N-gene rRT-PCR primers and probe.

### Cell lines for virus isolation

Cell lines used for the isolation of SARS-CoV-2 were obtained from the American Type Culture Collection (ATCC) and consisted of LLC-MK2 (Rhesus monkey kidney cells, catalog no. ATCC CCL-7) and Vero E6 cells (African green monkey kidney cells, catalog no. ATCC CRL-1586).

### Isolation of virus in cultured cells

Cells grown as monolayers in a T-25 flask (growing surface 25 cm^2^) were inoculated when they were at 80% of confluency. First, aliquots (100 µL) of the concentrated air sampler collection media were filtered through a sterile 0.45 µm pore-size PVDV syringe-tip filter to remove bacterial and fungal cells and spores. Next, the spent LLC-MK2 and Vero E6 cell culture medium was removed and replaced with 1 mL of cell culture medium, and the cells inoculated with 50 μL of cell filtrate. When virus-induced cytopathic effects (CPE) were evident, the presence of SARS-CoV-2 was determined by rRT-PCR.

### Quantification of SARS-CoV-2 genomes in sampled air

The number of viral genome equivalents present in each sample was estimated from the measured quantification cycle (Cq) values. To do so, a 6-log standard curve was run using 10-fold dilutions of a calibrated plasmid containing an insert of the SARS-CoV-2 N-gene that had been obtained from IDT Technologies, Inc. (Coralville, Iowa). The data was fit using equation (eq.) 1:

Eq. 1. y = (log10GE)(a) + b, where y = Cq value, a = slope of the regression line, log10GE is the base 10 log genome equivalents, and b is the intercept of the regression line.

### Sanger sequencing of SARS-CoV-2 genomes in material collected by air samplers

To obtain the virus consensus sequence prior to possible changes that might occur during isolation of the virus in cell cultures, a direct sequencing approach was used. Because the amount of virus present in the samples was low and thus unsuitable for common next-generation sequencing approaches, Sanger sequencing based on a gene-walking approach with over-lapping primers was used to obtain the virus sequence.^23^

### Next-generation sequencing the genome of SARS-CoV-2 isolated from NP swab

The vRNA extracted from virions in spent Vero E6 cell culture medium served as a template to generate a cDNA library using a NEBNext Ultra II RNA Library Prep kit (New England Biolabs, Inc.). Sequencing was performed on an Illumina MiSeq sequencer using a 600-cycle v3 MiSeq Reagent kit. Following the removal of host sequences (*Chlorocebus sabaeus*; GenBank assembly accession number GCA_000409795.2) using Kraken 2,^29^ *de novo* assembly of paired-end reads was performed in SPAdes v3.13.0 with default parameters.^30^

## Results

SARS-CoV-2 genomic RNA (vRNA) was detected by real-time reverse transcriptase quantitative polymerase chain reaction (rRT-qPCR) in material collected by air samplings 1-1, 1-3, 2-1, and 2-3, which had been performed without a HEPA filter covering the inlet tube. In contrast, in the presence of a HEPA filter, no SARS-CoV-2 genomes were detected in air samplings 1-2 and 2-2 (Table 1).

Virus-induced CPE were observed in LLC-MK2 and Vero E6 cells inoculated with material extruded from the NP specimen of patient 1 and from liquid collection media from air samples 1-1, 1-3, 2-1, and 2-3. Early CPE in both LLC-MK2 and Vero E6 cells consisted of the formation of cytoplasmic vacuoles that were apparent within 2 days post-inoculation (dpi) of the cells with material extruded from the NP swab and 4 to 6 dpi with aliquots of the liquid collection media from the air samplers. At later times (4 days onwards after inoculation of cell cultures with material from the NP swab, and 6 – 11 dpi of the cells with material collected by air samplers), rounding of the cells occurred in foci, followed by detachment of the cells from the growing surface. Some of the rounded cells detached in clumps, and occasional small syncytia with 3 -5 nuclei were observed. Apoptotic and necrotic cells were also observed. A representative collage showing the progressive development of CPE in Vero E6 cells inoculated with material collected during air sampling 1-1 is shown in Figure 2. Cytopathic effects were not observed and virus was not detected or isolated from the culture medium of samples 1-2 and 2-2, wherein HEPA filters had been affixed to the inlet nozzles of the air samplers, and were not observed in mock-inoculated cells which were maintained in parallel with the inoculated cell cultures.

**Figure 2.**
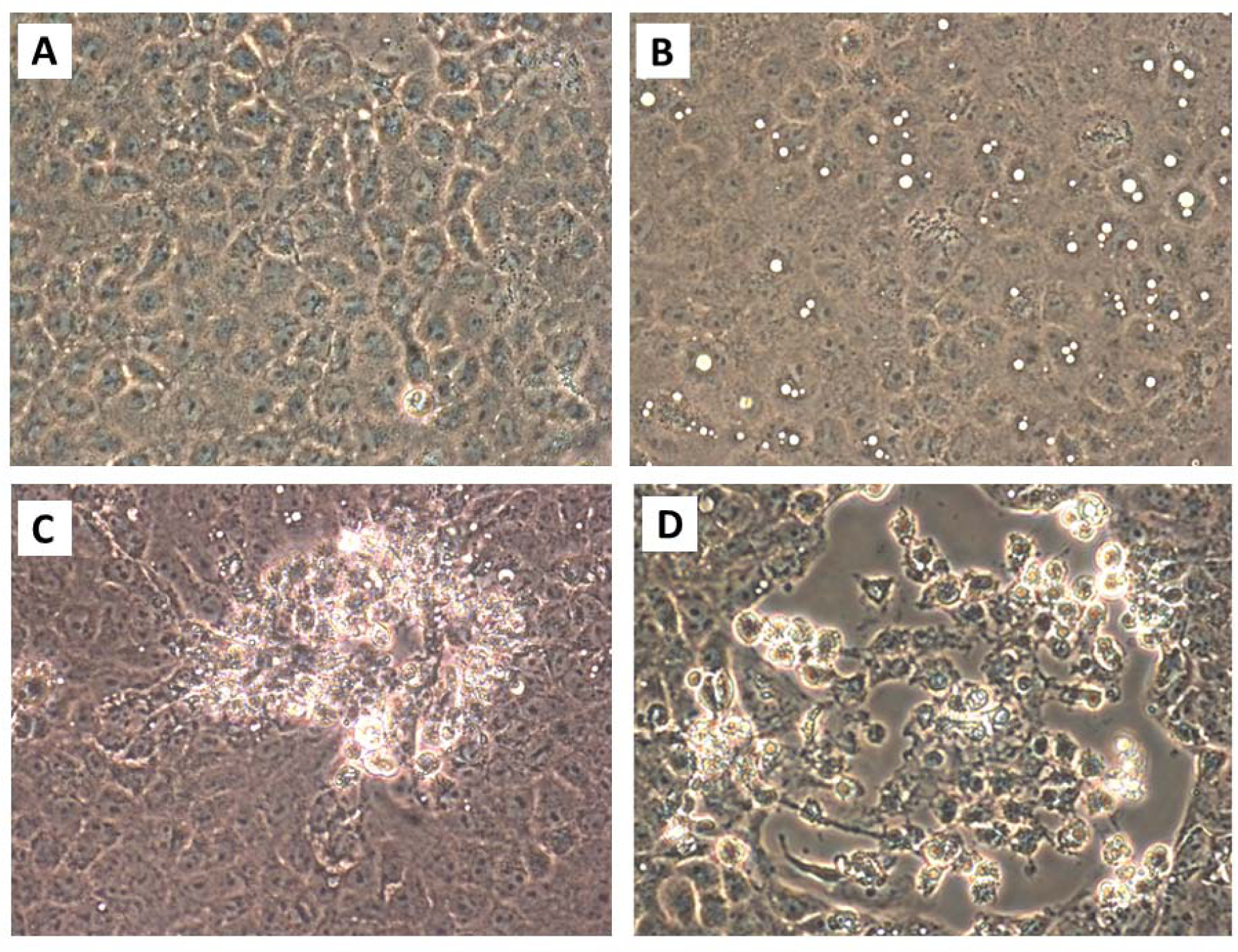
Cytopathic effects in Vero E6 cells inoculated with material collected from the air during air sampling 1-1. [A] Mock-infected Vero E6 cells, 10 days post-inoculation with sterile collection medium. [B]. Large cytoplasmic vacuoles in Vero E6 cells inoculated with collection medium from BioSpot sample 1-1 at 4 dpi. [C] Early focus of infection 7 dpi. [D] Focus of infection 10 dpi. Rounded cells that are detaching, some in clumps, are present. Attached cells remaining in this focus of infection have dark cytoplasms, some have large cytoplasmic inclusion bodies, and some cells are elongated. Original magnifications at 400X.

SARS-CoV-2-specific rRT-PCR tests were performed and the results indicated that the LLC-MK2 and Vero E6 cultures inoculated with collection media from air samplings 1-1, 1-3, 2-1, and 2-3 contained SARS-CoV-2 (data not shown). No other respiratory virus was identified in the samples using a BioFire FilmArray Respiratory 2 Panel (BioMérieux Inc., Durham, North Carolina), following the manufacturer’s instructions.

Whereas the concentration of SARS-CoV-2 genome equivalents per liter of air were estimated (Table 2), determination of the specific infectivity (ratio of SARS-CoV-2 genome equivalents present for every one able to infect a cell in culture) required performance of a plaque assay or a standard 50% endpoint dilution assay (TCID_50_ assay). Plaque assays could not be performed due to a nationwide non-availability of some critical media components (due to COVID-19 pandemic-related temporary lockdown of production facilities), so TCID_50_ assays were performed in Vero E6 cells to estimate the percentage of the collected virus particles that were viable. Estimates ranged from 2 to 74 TCID_50_ units/L of air (Table 3).

**Table 2.** Results of rRT-qPCR tests of materials collected by air samplers.

| Sample ID | Approx. distance (m) from head of patient 1<sup>b</sup> | Approx. distance (m) from head of patient 2<sup>b</sup> | rRT-qPCR test | Cq value | SARS-CoV-2 genome equivalents/25 µL rtRT-PCR test | SARS-CoV-2 genome equivalents/L of air |
| --- | --- | --- | --- | --- | --- | --- |
| 1-1 BioSpot | 2 | 4.6 | + | 36.02 | 2.82E+03 | 94 |
| 1-2 BioSpot + HEPA | 2 | 4.6 | - | - | - | - |
| 1-3 BioSpot | 2 | 0 (PD<sup>b</sup>) | + | 37.69 | 9.12E+02 | 30 |
| 2-1 VIVAS | 4.8 | 3 | + | 37.42 | 1.15E+03 | 44 |
| 2-2 VIVAS+ HEPA | 4.8 | 3 | - | - | - | - |
| 2-3 VIVAS | 4.8 | 0 (PD<sup>d</sup>) | + | 38.69 | 4.68E+02 | 16 |
| SARS-CoV-2 vRNA | N/A<sup>c</sup> | N/A | + | 29.53 | 2.20E+05 | N/A |
| N-gene<sup>a</sup> DNA control - 1 | N/A | N/A | + | 26.56 | 1.00E+06 | N/A |
| N-gene DNA control - 2 | N/A | N/A | + | 31.21 | 1.00E+05 | N/A |
| N-gene DNA control - 3 | N/A | N/A | + | 34.71 | 1.00E+04 | N/A |
| N-gene DNA control -4 | N/A | N/A | + | 37.74 | 1.00E+03 | N/A |
| N-gene DNA control - 5 | N/A | N/A | + | 40.41 | 1.00E+02 | N/A |
| N-gene DNA control - 6 | N/A | N/A | + | - | 1.00E+01 | N/A |
| Known positive (NP swab<sup>e</sup>) | N/A | N/A | + | 24.12 | 8.36E+06 | N/A |
| Negative (no RNA) control | N/A | N/A | N/A | - | 0 | N/A |

<sup>a</sup>N-gene, N-gene plasmid (positive control template).
<sup>b</sup>Distance from sampler inlet nozzle to patient's head.
<sup>c</sup>N/A, Not applicable.
<sup>d</sup>PD, patient discharged.
<sup>e</sup>NP, Nasal-pharyngeal swab from a person screened for SARS-CoV-2 at the UF EPI High-Throughput
COVID-19 Research Testing Facility.

**Table 3.**
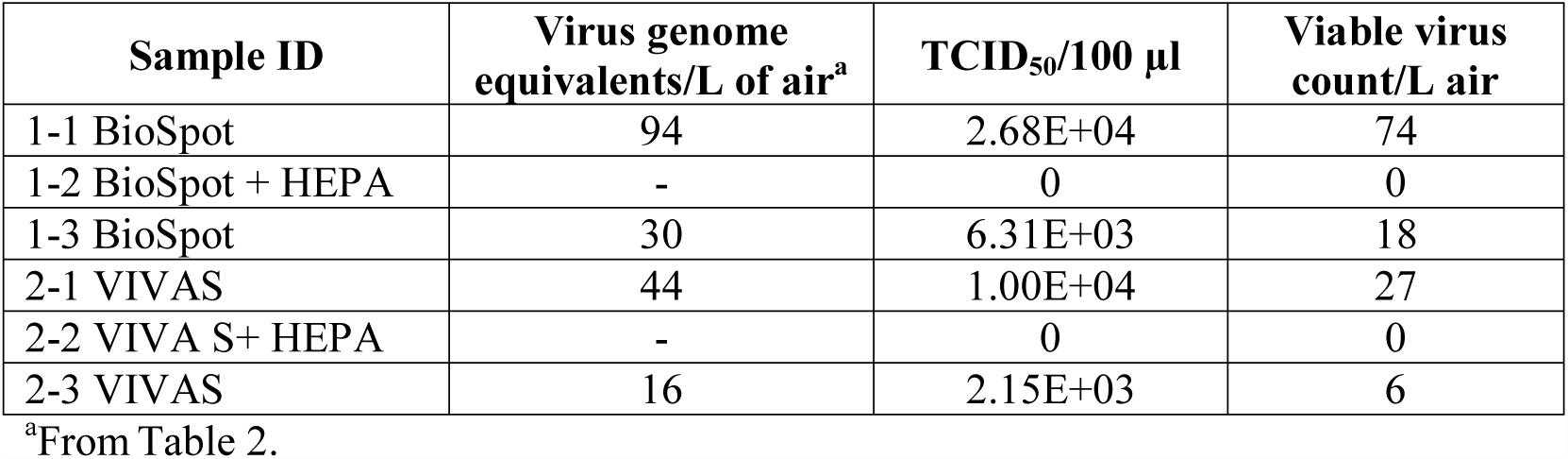
Estimate of viable virus counts based on TCID_50_ tests.

A nearly complete SARS-CoV-2 genome sequence was obtained by next-generation sequencing (NGS) of RNA purified from cell culture medium of Vero E6 cells 7 dpi with NP swab material from patient 1. The nearly complete genome sequence (and the virus isolate) were designated SARS-CoV-2/human/UF-19/2020, and this genome sequence has been deposited in GenBank (accession no. MT668716) and in GISAID (accession no. EPI_ISL_480349). Because the amount of virus RNA was below the threshold that could be easily sequenced by our NGS methods, Sanger sequencing was used to sequence SARS-CoV-2 RNA purified from the collection media of air samplers 1-1, 1-3, 2-1, and 2-3. One complete SARS-CoV-2 sequence was attained for RNA purified in the material collected by air sampling 1-1, and three nearly complete sequences for 1-3, 2-1, and 2-3, respectively. After alignment, comparisons of the three partial sequences with the complete sequence of SARS-CoV-2 in air sampling 1-1 indicated that the same consensus genome sequence were present in the virions that had been collected in all the air samplings. Moreover, they were an exact match with the corresponding sequences of the virus isolated from patient 1. This complete genome sequence of the virus collected by the air samplers (and the virus therein) were considered the same isolate and designated SARS-CoV-2/Environment/UF-20/2020, and this genome sequence has been deposited in GenBank (accession no. MT670008) and in GISAID (accession no. EPI_ISL_477163). The virus’ genomic sequence currently falls within GISAID clade B.1(GH), which is characterized by mutations C241T, C3037T, A23403G, G25563T, S-D614G, and NS3-Q57H relative to reference genome WIV04 (GenBank accession no. MN996528.1). As of 10 July 2020, SARS-CoV-2 clade B.1(GH) was the predominant virus lineage in circulation in the USA.

## Discussion

There are substantial epidemiologic data supporting the concept that SARS-CoV, which is highly related to SARS-CoV-2,^3^ was transmitted via an aerosol route.^31-33^ For SARS-CoV-2, there have also been two epidemiologic reports consistent with aerosol transmission.^15,34^ However, despite these reports, uncertainties remain about the relative importance of aerosol transmission of SARS-CoV-2, given that so far, only one study has provided weak evidence of virus isolation from material collected by air samplers.^21^ In other reports, attempts to isolate the virus were not successful. The current study takes advantage of a newer air sampling technology that operates using a water-vapor condensation mechanism, facilitating the likelihood of isolating the virus in tissue culture.

As reported in air sampling tests performed by others^9-11,21^ and in our previous report,^23^ airborne SARS-CoV-2 was present in a location with COVID-19 patients. The distance from the air-samplers to the patients (≥ 2 m) suggests that the virus was present in aerosols. Unlike previous studies, we have demonstrated the virus in aerosols can be viable, and this suggests that there is an inhalation risk for acquiring COVID-19 within the vicinity of people who emit the virus through expirations including coughs, sneezes, and speaking.

The amount of airborne virus detected per liter of air was small, and future studies should address (a) whether this is typical for COVID-19, (b) if this represented virus production relative to the phase of infection in the patient, (c) if this was a consequence of active air flow related to air exchanges within the room, (d) or if the low number of virus was due to technical difficulties in removing small airborne particles from the air.^26^

Our findings reveal that viable SARS-CoV-2 can be present in aerosols generated by a COVID-19 patient in a hospital room in the absence of an aerosol-generating procedure, and can thus serve as a source for transmission of the virus in this setting. Moreover, the public health implications are broad, especially as current best practices for limiting the spread of COVID-19 center on social distancing, wearing of face-coverings while in proximity to others and hand-washing. For aerosol-based transmission, measures such as physical distancing by 6 feet would not be helpful in an indoor setting, provide a false-sense of security and lead to exposures and outbreaks. With the current surges of cases, to help stem the COVID-19 pandemic, clear guidance on control measures against SARS-CoV-2 aerosols are needed, as recently voiced by other scientists.^35^

## Data Availability

SARS-CoV-2 genome sequence data are available in GenBank (accession numbers MT668716 and MT670008.

## Contributors

JAL, ML, ZHF, AJ, AEF, KC, JGM Jr, and C-YW conceived and designed the study. JAL, ML, KC, JG M Jr, and C-YW curated the data. JAL, ML, JGM Jr, and C-YW performed formal analyses of the data. JAL, ML, ZHF, AJ, JGM Jr. obtained funding for the work; JAL, TBT, MG, MU, SNS, KM, CJS, MMA, MAE, JCL, KS, and TBW performed experiments; JAL, M L, TBT, SNS, CJS, JCL, KS, TBW, JGM Jr, and C-YW established methods; JAL, ML, JCL, JGM Jr, and C-YW administered the project; JAL, ML, ZHF, AJ, JGM Jr, and C-YW provided resources; JAL, ML, JCL, JGM Jr, and C-YW supervised the project; JAL, JGM Jr., and C-Y Wu wrote the original manuscript draft; all authors revised the manuscript critically. All authors read and approved the final version of the manuscript.

## Acknowledgements

The authors thank Dr. Katherine Deliz (UF Environmental Engineering Sciences) for access to her laboratory for some essential engineering tasks, Drs. Christine Angelin and David Kaplan (UF Environmental Engineering Sciences) for providing critical supplies not readily available from vendors due to the COVID-19 pandemic, and Mark Dykes and Brad Pollitt of UF Health/Shands Hospital Facilities for providing room configuration and ventilation system info. Funding of work reported in this publication was partly supported by the National Science Foundation under Grant No. 2030844, partially by National Institute of Environmental Health Sciences of the National Institutes of Health award number 1R43ES030649, and funds made available by the UF Emerging Pathogens Institute and the Office of the Dean, UF College of Medicine. The contents are solely the responsibility of the authors and do not necessarily represent the official views of the National Science Foundation and the National Institutes of Health.

